# How just and fair are consent and debriefing for caesarean sections in Cameroon? An exploration of women’s and providers’ perspectives

**DOI:** 10.64898/2026.03.10.26348005

**Authors:** Jovanny Tsuala Fouogue, Veronique Filippi, Louise Tina Day, Mitsuaki Matsui, William Carter Djuatio Kenne, Bruno kenfack, Lenka Beňová, Miho Sato

## Abstract

Women-centeredness and a positive experience of caesarean sections (CS) care is contingent upon just, equitable and fair treatment of women during informed consent and post-operative debriefing, yet evidence from low-resource settings remains scarce. This study explored how routine practices of informed consent and post-operative debriefing for CS in West Cameroon speak to justice and fairness.

From March 2024 to August 2024, 69 purposively selected providers CS and 20 post-CS women were had face-to-face in-depth interviews. They were selected from the twenty hospitals in the West Region of Cameroon that recorded 100 or more CSs in 2022. Using an interpretivist paradigm, we generated codes inductively from verbatim transcripts and conducted thematic analysis. Our analysis drew on bioethical frameworks related to women’s rights (Fourie’s feminist approach to bioethics), epistemic and structural (in)justice (Fricker’s conception of epistemic injustice and Power’s and Faden’s work on power, advantage and human rights), and power relations (Odero’s work on patient - health provider power dynamics).

Consent procedure for CS was mainly verbal and conducted without protocols thus causing an informational deficit accentuated by the concealment or downplaying of risks and disproportionate involvement of third parties. Consent is generally initiated by the antenatal care or labour room midwife, confirmed by the attending physician and completed by the surgical theatre team (scrub and anaesthetist nurses). We identified several manifestations of injustice and unfairness in consent interactions comprising: a profound informational inadequacy contravening national medico-legal provisions; culturally hostile, informational deficient and disrespectful emergency consent routines; and unfairness towards rural women of low educational background. Routine debriefing practices were unfair towards low-income and unassertive women, the default being the absence of debriefing that was only provided upon woman’s resolute request. Four key barriers underpin these inequities: fear of CS, cultural misalignment, hospital structural constraints, and pervasive corruption.

Consent routines for both elective and emergency CSs in the West Region of Cameroon are characterised by several dimensions of injustice and unfairness that are not subsequently mitigated by post-operative debriefing which is by default absent. The underpinning of these are systemic drawbacks require redress to make consent, debriefing and CS at large more equitable, just and fair.

## Introduction

The technical provision of midwifery and obstetric care cannot holistically improve women’s physical, mental and social well-being without addressing their experience of care (1). In 2015, the World Health Organization (WHO) articulated a new vision of quality of care for pregnant women and newborn through a framework in which experience of care is on a par with technical provision of care (2). Based on this framework, the experiential dimension reposes on effective communication with women on their rights and the procedures being done, respectful and dignified clinical care and preferred social and emotional support. Over the past decade, global, regional and national health institutions have embarked on a journey to promote, adopt, and scale up respectful maternal care as a key component of the women-centredness domain of the quality-of-care framework (1,3,4). In a landmark statement in 2014, now widely endorsed across the globe, the WHO underscored the crucial need to implement respectful maternal and newborn care and generate data on both rights-based respectful and disrespectful care practices (5).

Caesarean section (CS) is central to emergency obstetric care and is the commonest major abdominal surgery worldwide. However, its associated mortality is 50 to 100 times higher in sub-Saharan Africa than in Europe and other high-income settings (6,7). While the clinical and technical dimensions of CS in African hospitals are well documented (8) (9,10) there remains a dearth of evidence regarding the experiential and interpersonal domains (communication, respect and emotional support), particularly concerning of informed consent and postoperative debriefing (3,11,12). Post-operative debriefing after CS is the information session(s) whereby the provider reports to the woman about the indication, procedures (including unforeseen), findings, eventual complications, implications for future reproductive life (i.e. inter-pregnancy interval, increased risk of CS, future place of delivery, workplace and domestic adjustments, management of possible complications etc.) and routine post-natal instructions (11,13).

In Cameroon, access to CS is low, with only 3.5% of women giving birth by CS in 2018 (14). Besides, the technical quality of this procedure is poor with evidence showing high prevalence of post-surgical complications (14–17). There is little evidence on informed consent, postoperative debriefing, and respectful CS care in the country (18,19). The country is a party to most, if not all, United Nations and African Union legal instruments to promote and protect women’s rights (20,21). Several national laws aim to promote, protect, and restore women’s welfare and dignity. The Constitution sets out that “every person has a right to humane treatment in all circumstances” and that “the Nation shall protect women” (22). Through its Law No. 96/03 of 4 January 1996, which lays down an outline law in the health domain, the State committed itself to ensuring “access to integrated and quality health care”. The Cameroon Code of Medical Ethics instructs physicians to “always treat patients with politeness and correctness” and to “seek their consent in all circumstances” (23). Furthermore, “physicians owe their patients truthful, accessible, and relevant information on their medical conditions and proposed treatment throughout the episode of illness” (23). Yet there are widespread violations of patients’ rights in health facilities across the country and regular controversies related to poor quality of CS care (20,24–27).

Quality CS requires documented verbal or written informed consent from the woman to meet the exigencies of biomedical ethics, in which justice is a moral principle alongside respect for autonomy, beneficence, and non-maleficence (28). Justice is defined as “a group of norms for fairly distributing benefits, risks, and costs” (28). As such, justice in healthcare centers on the systematic application of established laws (e.g. codes of medical ethics) and rules (e.g. clinical practice guidelines) to deliver clinical services that are rightfully due to patients. Justice and fairness both relate to the notion of treating one as they deserves; but while justice is rule-based and related to established standards of rightness regardless of the specificities of particular cases, fairness, on the other hand, focuses on individual equity and proportionality (29). Fair provision of healthcare factors in patients’ subjective needs, individual circumstances, and contexts to deliver equitable and unbiased services. Fricker defined epistemic injustice as “a wrong done to someone specifically in their capacity as a knower” (30). It has two facets: testimonial injustice and hermeneutical injustice. While testimonial injustice is inflicted because the victim is denied the necessary credibility to validate their status of knowers (participatory prejudice) or their knowledge (informational prejudice), hermeneutical injustice occurs because perpetrators are unaware of the knowledge at stake (30–32). Powers and Faden furthermore theorised structural injustice as a defining feature of institutional arrangements and the web of social norms in which people live (33). In their theory, human rights violations, disadvantage, and unfair power relations mutually reinforce each other to produce injustice. Hermeneutical and testimonial epistemic injustices in obstetric care are illustrated by the discount of the pain testimony of Black and Latina American women based on fallacious racial essentialism (beliefs in higher physiological resistance to pain), leading to profound distributive injustices in analgesic treatment during childbirth and immediate post-partum (34,35). Regarding structural injustice, in 2016, Kidd reported on how the predominance of biomedical approach of clinical care, the efficiency and profit orientation of many modern health systems feed the hermeneutical stance of care providers (36). This was acknowledged in the United States of America, where the section “Psychological, Social, and Biological Foundations of Behaviour” was added to the Medical College Admission Test in 2015 to ensure that future health professionals are culturally competent in order to offer patients a positive experience of care thereby preventing epistemic injustices (37).

Given the dearth of evidence to inform actions to improve women’s experience of care in Cameroon, we aimed to explore how routine practices of informed consent and debriefing for CS speak to justice and fairness. Our analysis draws on bioethical frameworks related to women’s rights (Fourie’s feminist approach to bioethics), epistemic and structural (in)justice (Fricker’s conception of epistemic injustice and Power’s and Faden’s work on power, advantage and human rights), and power relations (Odero’s work on patient - health provider power dynamics) (30,33,38,39).

## Materials and methods

### Qualitative approach, guiding theory, and research paradigm

This qualitative research used in-depth interviews (IDIs) followed by thematic analysis of CS providers’ (thereafter referred as providers) views and women’s lived experiences. IDIs were chosen to explore women’s experiences of consent and debriefing for CS because of the anticipated deep dive into their intimate health records and private lives. Providers were also interviewed individually because they formed a diverse group of specialized professionals working in several units with interdisciplinary and hierarchical relationships. We deemed IDIs appropriate to probe details in each speciality and avoid the effect of professional power dynamics on free speech. Moreover, conflicts between clinical specialities and units that potentially affect consent and debriefing were expected to be freely recounted during the IDIs. Naturalistic observation was considered for data collection. However, it would have been challenging for planned CSs, given that consent discussions in such cases typically occur during multiple antenatal consultations. Likewise, observing consent and debriefing interactions for a single emergency CS in each study hospital could have been biased by the Hawthorne effect, amplified by the necessary clinical background of the observer.

This study adopted an interpretivist approach. Interpretivism is a research philosophy that upholds that truth and knowledge are subjective, as well as historically and culturally situated, based on people’s lived experiences and understanding (40). It emphasises individual agency and interpretation within social structures and institutions. Participants’ lived experiences and interpreted perspectives on routine practices of consent and debriefing were appraised within the context against the backdrop of the dimensions of the four bioethical frameworks we mentioned in the introduction.

### Study design

#### Study sites and context

We included 20 hospitals in the West Region that recorded at least 100 CS in 2022 spanning nine health districts. The West Region is one of the ten Regions of Cameroon (41,42). Its population size was 2,398,224 inhabitants in 2024 and it suffers a persistent severe shortage of human resources for health (43–45). Almost all deliveries (96.9%) in 2018 were facility-based, of which 10% were through CS, mainly in emergency contexts (80%) (46,47). Household direct out-of-pocket fees for CS remain prohibitive and caused catastrophic health expenditures in 42% of cases in two neighbouring regions in 2023 (48). Both the literacy rate and Gender Inequality Index are high, and patriarchy is deeply rooted in the collectivist social fabric of communities (49,50).

#### Participant selection

We purposively included two categories of participants: providers and women within 30 days of their CS (hereafter referred to as post-CS women), along with their accompanying relatives. We invited post-CS women by phone and conducted IDIs at locations of their choice outside hospitals. Every post-CS woman who reported limited involvement in consent interactions (due to an emergency procedure or the dominant role of a third party) was asked to indicate the relative who accompanied her for the CS. We then included these third parties in the IDIs after consent. We included participants until the sample size met data saturation and exhaustive coverage of the 20 hospitals (51,52).

#### Data collection

Drawing on FIGO guidelines on ethics, surgical consent and respectful care, the Cameroon national code of medical ethics, the international code of medical ethics, African guidelines on post-CS care, guidelines opinions of obstetricians-gynaecologists working the West Region, JTF (a male Obstetrician Gynaecologist) and WCDK (a male social scientist) designed the interview guides that were reviewed by all co-authors (11,23,53–56). After pretesting the guides, they conducted face-to-face IDIs between 1^st^ March 2024 and 30^th^ August 2024. The IDIs with providers were conducted in private in hospitals offices either in English or French depending on their preferences. The IDIs with women were conducted at their homes in the presence of a female community health worker to ensure contextual sensitivity. We audio-recorded the IDIs, took field notes and did not any of the IDIs.

### Data analysis

We (JTF and WCDK) inductively developed the codes using NVivo® 14 (Lumivero LLC). After coding the first four transcripts of each interviewee category, we merged the two individual codebooks. To enhance trustworthiness, we constantly memoed, systematically referred to field notes, and compared our codes across data sources. In addition, we triangulated information between the accounts of providers from the same hospitals and between women’s and providers’ reports. Saturation was reached with respect to the proceedings (processes, actors, and contents) of consent and debriefing after coding the transcripts of 16 IDIs with post-CS women and providers from 12 hospitals. However, new meanings related to contextual sociocultural features kept emerging, warranting continued data collection and coding for all the remaining hospitals (51,52). JTF edited the harmonised codebook that was further amended and approved by the larger research team (VF, MS, LTD and MM).

Building on the final codebook, we generated cascades of informed consent and debriefing and elaborated initial themes delineating (un)fairness to women, (dis)respect for their rights under the prevailing normative framework, (as)symmetrical power relations, and epistemic and structural (in)justice through the analysis of emerging patterns and identification of relationships (contradiction, similarity, causality, hierarchy, co-occurrence, and intersection). We then fine-tuned and reported the results in accordance with The Consolidated guidelines for REporting Qualitative research (COREQ) (57,58).

### Ethics

This study was approved by the Institutional Review Board of the London School of Hygiene & Tropical Medicine (Reference: 29898) and the Regional Ethics Committee for Human Health Research for the West Region (Reference: N° 984/25/10/2023/CE/ CRERSH-OU/VP). This study was conducted in accordance with the provisions of the Helsinki Declaration (2013) and relevant Cameroonian regulations.

## Results

All the invited providers participated, while two of the 22 post-CS women declined, one for personal reasons and the other because of relocation to another Region. Thus, we conducted 69 IDIs with providers and 20 with post-CS women. Half of the IDIs with post-CS women included accompanying relatives who were often female, upon the woman’s request. No participants dropped out during the data collection.

### Participants’ characteristics

The median age (range) was 38 (23–59) years for providers and 28.5 (17–42) years for post-CS women. Half of the providers were female, 40% worked in maternity wards, and 60% worked in surgical units. Seventy-five percent of post-CS women were in marital union, 65% were Christian, and 80% had experienced an emergency CS (Table 1).

**Table 1.**
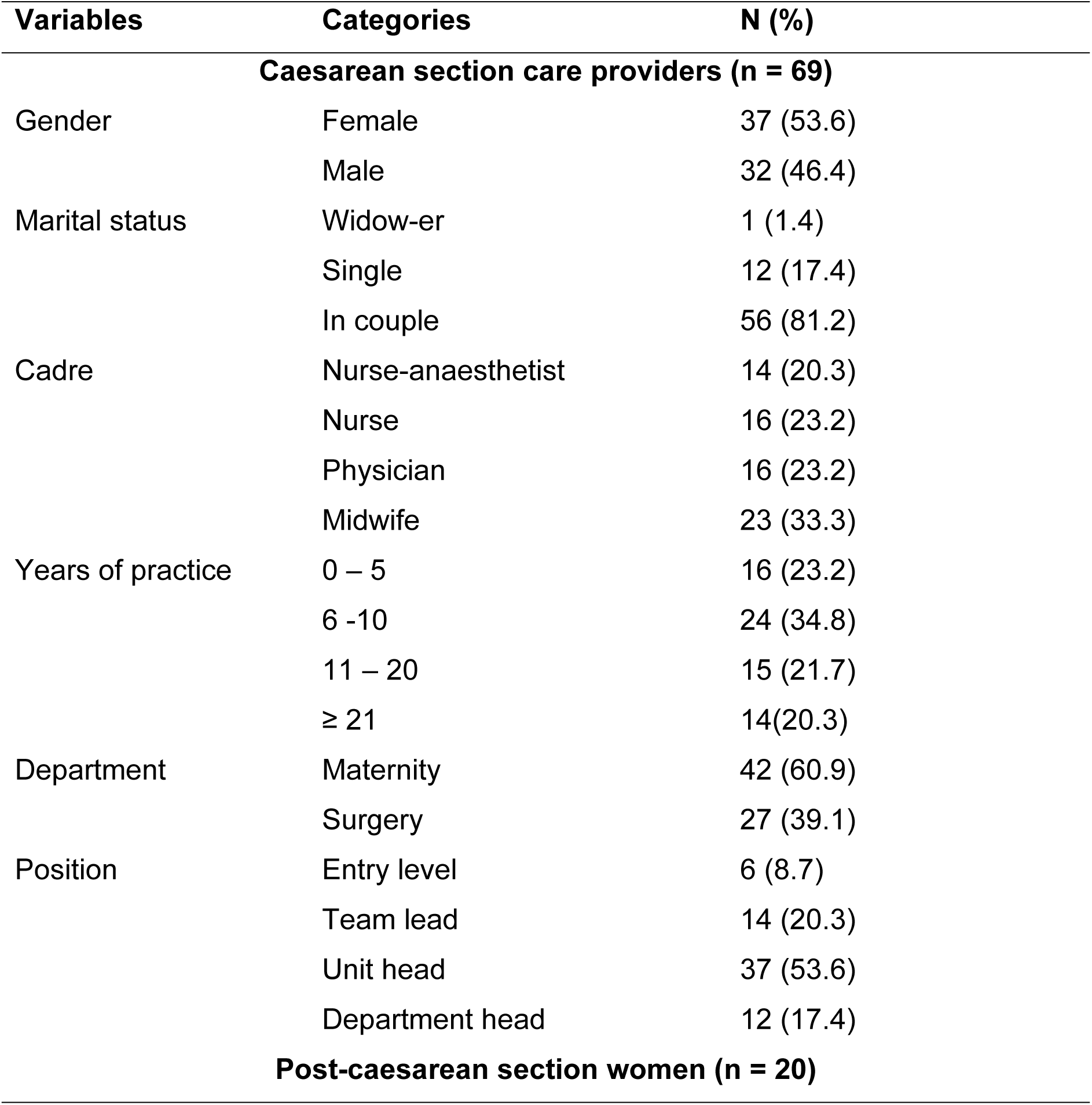
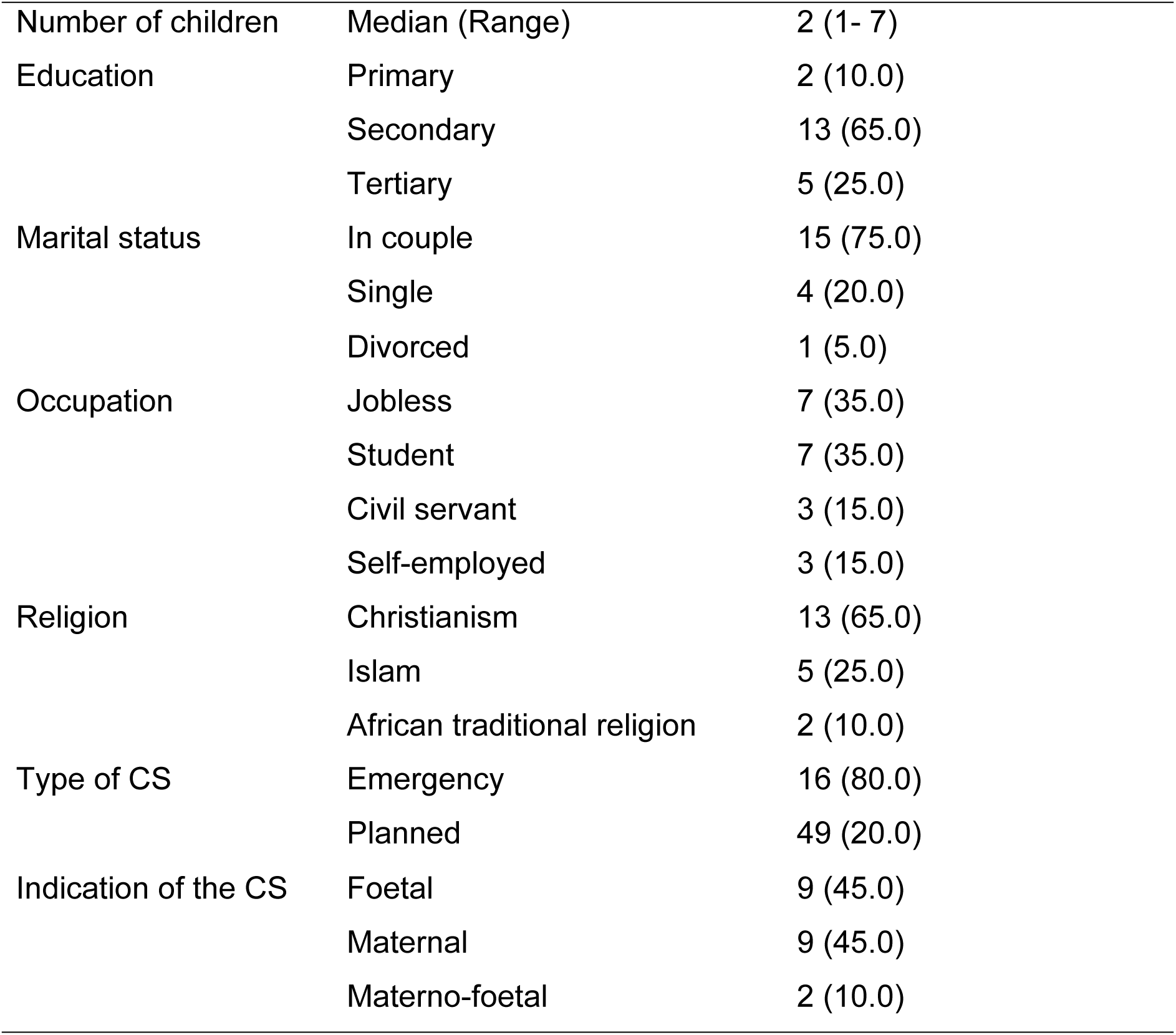
Participants’ characteristics. How just and fair are consent and debriefing for caesarean sections in Cameroon? An exploration of women’s and providers’ perspectives from 20 twenty hospitals in West Cameroon. March – August 2024.

CS: caesarean section; IDI: In-depth interview

### Routine practice of consent for caesarean section

Procedures for emergency and elective CS were different and are presented below in terms of specific cascades of events, venue and actors (Fig 1).

**Figure 1.**
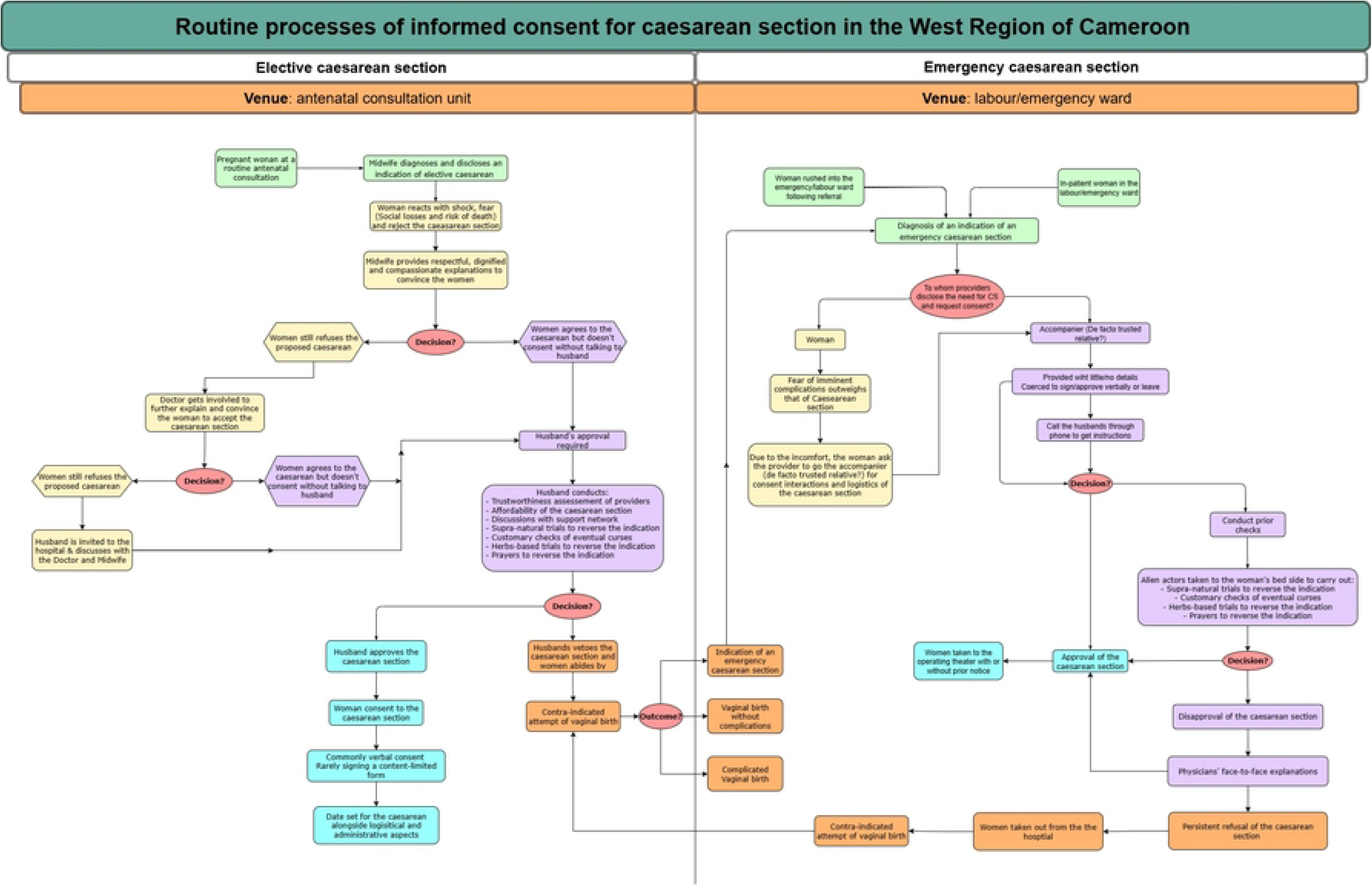
Routine processes for obtaining informed consent for emergency and elective caesarean sections in the West Region of Cameroon. March 2024 – August 2024.

#### Consent for elective caesarean section

The cascade of interactions (Figure 1) for elective consent begins during antenatal care, when an indication for CS is identified by the midwife, confirmed by the physician, and communicated to the woman. Fear, anxiety, and refusal are the default responses of pregnant women. After efforts to provide painstaking, respectful, and compassionate explanations, often during several antenatal visits, the midwife usually obtains the woman’s verbal approval but not their final consent which is generally subject to the husband’s clearance.

> *“During the antenatal consultation, we must make the woman understand that a CS is not the end, it not a sanction because for many women when we talk of CS, it is like if the sky was falling on their heads, like if we were sanctioning them”. **A Midwife***.

> *“For elective CS, we usually have much time to give details to the woman herself; some come with their husbands, mothers or mothers-in-law.” **A general practitioner***.

> *“Upon the doctor’s mention of the CS, I started an uncontrollable outcry. Despite his admonitions to remain calm, I had already commenced a mourning ritual of weeping. He comforted me, asserting that the procedure was not a funeral and that lamentations and such displays of grief were detrimental within the surgical theatre”. **A post CS-woman***

#### Consent for emergency caesarean section

*“Given that we don’t have a consent form, we endeavour to explain.” **A Midwife***

For emergency CS, the consent cascade begins when a midwife diagnoses the need for an emergency CS during or before labour, obtains the physician’s confirmation, and informs the woman. Classically, the latter, out of fear and stress, precipitously approves the CS to stop suffering caused by labour pains or pregnancy complications and asks the midwife to engage with her accompanying relative, usually someone she trusts. In some cases, a minority of midwives skipped the woman and directly engaged with the accompanying relative, most often the husband to secure consent, as indicated by two post-CS women.

> *“They did not even announce me* [the CS] *… I noticed they would do the CS because I saw they placed the urinary catheter; they undressed me and took me to the operating room.”*

> *… “It is not normal: they do a CS, and they don’t inform you; they just prepare you and take you to the operating room.” **A post-CS woman***

> *“For the operation, I was surprised. In the operating room, they told me they had already agreed it with my husband. I don’t know why they didn’t tell me beforehand. … I don’t know what their intention was in keeping it from me. But when I arrived, I found myself ambushed like that; they told me: ‘Your husband has agreed.’ But I asked, “why am I being operated?”” **A post-CS woman***

Women who steadfastly refused CS, despite being provided with information and the involvement of their accompanying relatives, were either forcibly referred to higher-level hospitals or turned away.

> *“Well, we answer this: if you don’t want that the surgery* [caesarean section] *to be done here* [the index hospital], *take her elsewhere”. **An anaesthetist nurse***

> *“They* [health care providers] *said we must go to the operating room and that if I accept, they will operate on her, but if I refuse, they will refer her to another hospital. It was only then that I signed.*” ***An accompanying relative (Her mother)***

> *“We insisted that let us go and operate. This woman refused,* [preferring] *that we should do everything so that it* [the baby] *should come out vaginally. I said no; I cannot take the risk because I see that I will not be able to take out this child vaginally. Finally, we did not agree, and I had to say* [to the woman] *go the District hospital.” **A general practitioner***

### Inadequate provision of information to women before and after the caesarean section

The provision of relevant and comprehensible information to women and accompanying relatives while seeking consent for CS was quite limited, despite good clinical practice guidelines and legal requirements in Cameroon. Post-CS women and their accompanying relatives complained that during emergency consent interactions, access to information on the proposed CS was profoundly hampered by providers’ reluctance to answer questions.

> Interviewer: *Did the care providers made sure you read the form you signed?*

> An accompanying relative: [The provider] *simply told me to sign, write my identity card number, and we went to the operating room*.

Furthermore, providers working in the labour rooms insisted that seeking emergency consent should be done exclusively by their colleagues working in the operating rooms (surgeons, operating room nurses and anaesthetist nurses), who are exclusively liable for the procedure and are best placed to accurately answer the woman’s questions. This reveals that incomplete information was routinely provided to women by the labour room staff who felt unentitled to do so. Disclosure of the potential harm caused by CS was skipped or lightly touched by midwives in the labour rooms and surgeons providers who admitted to actively concealing the risks of the procedure during consent interactions.

> “Women are often surprised when we talk about the risks of a caesarean section. This is because health workers do not usually explain these complications to them.” **A General Practitioner.**

> “Interviewer: *Did they tell you about the risks* [of the CS]*?* - A post-CS woman: *No!”*

Both post-CS women and providers from most hospitals indicated that debriefing was not routinely done, thereby depriving women of due information. When assertive post-CS women requested accounts of the procedure, answers were limited and often not truthful regarding the eventual complications.

> *“After my C-section, I did not receive any medical supportive communication. I had to ask many questions and lobby with my connections just to get a little information. No midwife or doctor sat down with me to explain how to recover after what I went through. I was left without any proper guidance.”* **A post-CS woman**

### Culturally hostile, informationally deficient and disrespectful consent for emergency caesarean sections

Women undergoing emergency CSs received rushed consent processes characterized by inadequate information disclosure and a profound disregard for their cultural specificities; this was particularly pronounced for unassertive rural women with a low educational background.

#### Consent for emergency caesarean sections

> *From the midwifery perspective, “Generally, it’s extremely frustrating, because we have cases where there’s a real emergency, and what the husband decides to do is go look for a traditional healer. Or when there’s a malpresentation, and they tell you a traditional healer will give some medicine that will turn the baby inside the womb. We see cases like this very frequently.” **A Midwife***

> *“When complications happen to women while in the hospital, it is our* [hospital staff] *responsibility. So, the traditional healer who interferes in the emergency CS consent interactions has no responsibility for that. This is why we are strongly against what they [traditional healer] come and do to women in labour” **A Midwife***

The content of informed consent interactions differed significantly for women undergoing emergency CS across all hospital settings in terms of cultural friendliness, disclosed information, and tact. Providers underscored their hostility towards commonly practiced spiritual, traditional, and cultural rituals (scarifications, topical ointments, ingestion of herbal decoctions and mastication of medical barks) by women and their accompanying relatives before emergency CS. They reported obstructing these rituals as much as possible, arguing that they disrupt the continuum of care and increase the risks of both surgery and anaesthesia. Furthermore, providers justified providing less information on CS to avoid delaying urgent procedures. In contrast, as described above, women undergoing elective CS benefited from several rounds of explanations by the attending providers and had sufficient time to perform rituals.

Unlike women undergoing elective CS, those undergoing emergency procedures were not always treated with respect during consent interactions. They reported many instances of overt mistreatment, such as verbal abuse and precipitation in life-threatening circumstances, where they needed considerable attention and support.

> *“The way they talk to women in distress there,* [hospital] *is not easy”. **A post-CS woman***.

#### Consent unfairness towards rural women with low educational background

Unfairness toward rural and less educated women in the CS consent process was evidenced by the absence of an adaptive communication approach, such as translating information into local languages or simplifying complex medical terms, necessary to support their decision-making capacity. More educated urban women themselves recognised this inequity and pleaded that providers should proactively centre communication on less educated and subservient women who tend to rely on their relatives as proxy consenters.

> *“Clinicians must calibrate their communication to the low health literacy levels of less-educated rural women, while remains sensitive to the profound influence of marital deference on their autonomy.” **An urban highly educated post-CS woman.***

Providers acknowledged that even in lower-level healthcare facilities, where most staff members spoke the local language, their workload prevented them from explaining the minimum informational content of the consent form. In contrast, staff in higher-level health facilities struggled to translate consent related information into local languages for women and accompaniers from low educational background.

***A General Practitioner:*** *Usually accompaniers are elderly mothers, and same as the women, they don’t really understand official language and what is happening*.

***Interviewer:*** *I see*.

***A General Practitioner:*** *If we really want the message to go around, we need somebody who has a minimum level of education*.

***Interviewer:*** *And commonly it is the husband?*

***A General Practitioner:*** *It is the husband, yes*.

***Interviewer:*** *And when the husband is not around?*

***A General Practitioner:*** *He will be contacted on phone*.

***Investigator:*** *What do you do when the husband is not reachable?*

***A General Practitioner:*** *Now we will find a third party to translate for the grandmother.”*

Most providers reported that rural women’s over-reliance on ancestral medicine delivered by traditional doctors, spiritual healers and herbalists intensifies the clash between knowledge systems during consent transactions with providers spearheading the hospital hostility toward indigenous healing practices.

> *” In villages and countryside, women use traditional healers (marabouts) much more than in cities. Before coming to the hospital, many women visit these healers who give them false hope. Sometimes, the healers even come with them. In the city, it is harder to find these people, so women are less influenced by these traditional beliefs“* **A midwife**

### Unfair debriefing practices towards low-income and unassertive women

In the few hospitals that delivered post-CS debriefing, it did not align with the values and real-life requirements of post-CS women who deplored the lack of supportive communication towards a conducive household environment for recovery and birth spacing.

> *“Yes. Normally when I was getting out* [of the hospital]*, they* [health care providers] *should have told me that: at home, you must not do this, you must not eat this kind of food to avoid having problems.” **A post-CS woman***

In addition, unless they paid, low-income women were at a higher risk of neglect during post-CS care, including debriefing. Indeed, providers admitted that demotivation due to their low and irregular wages and often poor work environment (work overload, absence of incentives, and stockouts of supplies) made them vulnerable to increased attention for post-CS women consenting to under-the-table payments or gratuities.

> *“She* [the attending nurse] *firmly says that if we don’t buy the gloves from her, she will not administer the medicines, because she would have received 750 XAF* [1 pound sterling]*. (laughter).” **A post-CS woman***

Providers further indicated a substantial risk of double-standard care during debriefing, which requires the attention of hospital managers and policymakers. This was supported by the fact that women and their kin decried numerous instances where post-CS care was contingent upon illicit surcharges and informal financial demands. The impact of Providers’ job dissatisfaction is well known. Consequently, many women and their husbands described how they mitigated this by spontaneously tipping or bribing healthcare providers.

> *“You will see that people* [health care providers] *are always upset when they interact with people* [women] *who don’t pay cash for the care services.” **A general practitioner***

> *“When I enter a hospital, knowing how underpaid the staff are, I prefer to motivate* [tip] *from the start, so it does not affect the way we are treated.” **An accompanying relative (husband)***

Most providers and post-CS women acknowledged the absence of systematic debriefing in public hospitals. Apart from clinical care, bill settlement is the only element checked before post-CS discharge home with an appointment for the sixth-week postnatal consultation. Substantial unanticipated extra costs were reported as a cause of discontent for women and their support networks, who blamed the unpredictability of the total fees. They added that in those hospitals, the only way to receive debriefing was to request it assertively, and yet it was commonly done by other providers rather than by those who had conducted the CS.

> *“In that hospital, if you don’t ask questions they* [doctors and midwives] *will not explain you anything about your cesarean section. Apart from injections and tablets, they just make sure you pay your bill before you leave.” **A post-CS woman***

## Discussion

### Main findings

The informed consent process in Cameroon was reported to differs between emergency and elective CSs. In elective situations, interactions were described as more woman-centred and respectful. Conversely, in emergency situations, beyond the constraints imposed by time, unfair and unjust consent interactions took the form of limited involvement of the woman in decision-making, limited information, many instances of mistreatment, and culturally unfriendly attitudes. Moreover, the documentation of consent was inadequate in some cases. A further salient injustice concerned post-operative debriefing with women: its near absence was largely normalised, with limited attention to woman’s needs, alongside unfair treatment of poorer and non-assertive women.

### Findings in context

The inadequate quality of consent for emergency CS has been reported in countries across income categories (19,59–63). In our study, in Cameroon, this comprised a blatant violation of women’s right to information that was worsened by the near absence of postoperative debriefing, which is an opportunity to inform women post-operatively (23,56). Given that common delays in emergency referrals for CS do not leave enough time for in-depth consent interactions and that birth plans set out during antenatal consultations often lack sufficient details on unforeseen CS, debriefing presents an ultimate opportunity for Providers to catch up on their legal duty of disclosing information. Unfortunately, the lack of standard operating procedures to streamline the practice of post-CS debriefing in public hospitals is a common feature of postnatal care in sub-Saharan Africa (11,64). In Cameroon, skipping post-CS debriefing, as is the case in most hospitals in the West Region is unlawful.

Compared to women undergoing elective CSs, who received adequate information in responsive communicative interactions, those undergoing emergency procedures experienced unfair consent practices that did not account for their emergency-related needs. Beyond this, organizational injustice manifested in late referrals for emergency CSs, providers’ job demotivation, and poor working conditions. These structural constraints exposed poorer women to further unequal treatment during debriefing because they were unable to provide informal payments, bribe or tip. This income-based injustice appears to be characteristic of Cameroon’s health system and has been reported in African countries with similar levels of corruption(62,65,66).

The patterns of unfairness described above align with Powers and Faden’s theory of structural injustice (33). Specifically, four of the five core elements of well-being identified in their framework are directly eroded by structural injustices in the delivery of consent and debriefing: health (of the woman-baby dyad), knowledge and understanding (of the CS procedure), and equal respect and self-determination (within consent and debriefing interactions). Moreover, as we shall illustrate, all these causative unfair conditions are embedded in the institutional fabric of hospitals.

First, there is a major asymmetry of power between the woman and the Provider during emergency consent on the one hand and between the woman and her accompanying relative standing on behalf of a male relative (husband or father). This is due to the intertwining of testimonial and hermeneutical epistemic injustices (30).

With respect to testimonial epistemic injustice, women suffer identity injustice underpinned by gender and marital norms which subordinate them to men (husbands or fathers); consequently, there is a prejudice deficit of credibility from healthcare providers who align with those norms.

Simultaneously, hermeneutical epistemic injustice manifests in the collective deficit of the interpretive abilities of care providers as a specific group which is unable to make sense of the unfair disadvantage women experience during CS-related consent and debriefing. This is a second-order barrier to any attempt by women to communicate their disadvantages and claim fairness.

Evidence in Tanzania indicates that tackling this issue is within the reach of public hospital managers, thereby indicating leadership shortcomings in public hospitals, all the more so as such complaints were not raised in private faith-based hospitals in the West Region (3,18,67). It is worth noting that the above-mentioned injustices in consent and debriefing practices, namely non-consented CS, corruption, and discriminatory care, are all recognized components of obstetric violence (68).

Notably, our findings reveal a striking congruence between providers’ reluctance to disclose the risks of CS and previously substantiated women’s widespread preference in the West Region not to be informed about these risks as well as their fear (69). Likewise, a study in the United Kingdom and another in Malawi reported that little information on the risks of CS was disclosed to women (61,70). Women’s great fear of surgical or anaesthetic complications is part of pre-CS anxiety which is a worldwide phenomenon more pronounced in high maternal mortality regions (62,70–78). In a systematic review on patients’ and providers’ experience of consent, Convie et al. found that patients’ fear and anxiety influenced their engagement with the informed consent process (12) . In Nigeria, Agu et al. found that less than half of the healthy adults from three communities were willing to be told about complications and would change their surgeon who told them about complications during consent discussions (12,79). Though women seem to have normalized the non-disclosure of CS risks during consent discussions, providers should strike a balance between this unspoken expectation and good clinical practice of clearly disclosing the risks of any surgical procedure by toning down the complications in a compassionate manner, using interpersonal communication techniques to inform without amplifying fear.

Individual-related injustice came into the picture when some CS providers verbally abused and blamed them for risky behaviours that led to emergency CSs or unlawfully denied them further care upon informed refusal of the proposed CS. The latter is probably underpinned by providers’ reluctance to have maternal deaths on their shoulders resulting from women’s refusal.

Alongside WHO guidelines, legally binding instruments from the United Nations, the African Union, and Cameroon’s legislative armamentarium set out that, regardless of the circumstances, women deserve to be treated with respect and courtesy in healthcare settings, and providers ought to be proficient in interpersonal communication (3,23,80). In the same vein, it is a serious offense to drive a woman away from a hospital because consent to perform a CS is not granted, as reported by the providers. Indeed, both the World Medical Association’s and Cameroon’s codes of medical ethics state that physicians should inform, document, and respect the refusal of CS and continue to attend to the women (23,56). This practice which reflects either ignorance of the law or impunity of the providers, corroborates the widely decried malpractices and abuses perpetrated against patients in health facilities across Cameroon (20,21).

Cultural epistemic injustice in consent and debriefing interactions in most of our study hospitals was reflected in the flat rejection by providers of the conduct of metaphysical customary and traditional rituals by women and their relatives before emergency CSs. Given that the essence of those rituals inspired by indigenous knowledge makes up the core of women’s social identities, providers could be perceived as epistemic oppressors who exclude the indigenous contribution to the healing enterprise, thereby trimming the acceptability of the procedure and trust in doctors and hospitals (81). Worse, the fallacy of their arguments that the rituals are totally incompatible with safe CS was demonstrated by the inclusive attitude of the minority of providers who examined the various rituals and allowed those deemed harmless. The hermeneutical stance adopted by CS providers does not only contradicts the women-centred maternity care, which promotes culturally respectful practices aligned with women’s beliefs and values, but also reflects a minimization of the feminist bioethical critique of the androcentric approach to healthcare (3,38). Indeed, from a decolonial perspective, even a universally proven biomedical “global” or “western-imported” procedure like CS should be deployed in local (former colonial) contexts with a pluralistic and inclusive mindset that values indigenous health practices and engages with those deemed “compatible” (82) (83).

### Strengths and limitations

The credibility of these descriptions of consent and debriefing practices in Cameroon is strengthened by the inclusion of hospitals across all levels in nearly half of the health districts of the West Region, and by triangulation of data from providers, women, and their accompanying relatives. Finally, these features enhance the transferability of our findings to similar settings.

The main limitation of this study is the lack of direct observation of consent and debriefing procedures, which could have generated more detailed insights on justice and fairness.

### Policy, practice, and research implications

The design, adoption and maintenance of a context-relevant standard operating procedure for consent and debriefing could address the major injustices and unfairness uncovered by this study. Unjust informational deficit during consent and debriefing for CS could be addressed by the systematic use of a predefined consent form and a post-CS discharge checklist. The training offered to providers prior to the introduction of the previous documents would help tackling disrespectful and culturally unfriendly practices by encompassing modules on compassionate care and women’s positive experience of hospital services. Last, a rapid scaling of the ongoing piloting of universal health coverage scheme in the West Region would stop the blatant distributive injustices towards vulnerable women (rural, low-income and low educational background).

Further research should focus on the co-design and evaluation of such a standard operating procedures for informed consent and debriefing for CS.

## Conclusion

Elective and emergency caesarean sections in the West Region of Cameroon are characterised by different dimensions of fairness and injustice. However, emergency CSs are particularly affected by inadequate information provision and limited capacity or willingness to respond to women’s cultural expectations. These shortcomings are not mitigated by post-operative debriefing, which is often absent or insufficient. Four key barriers underpin these inequities: cultural clashes, structural constraints at the facility level, pervasive corruption, and fear of caesarean section. All require attention if consent practices are to become more equitable, just and fair.

## Data Availability

Data related to this research are available from the corresponding authors upon reasonable request.

## Acknowledgments

We thank all the participants who generously lent their time to this research. We are also immensely grateful to Mr. Etienne Wado (Regional Sexual and Reproductive Focal Officer) and to Dr Alain Patrick Kamleu Tchatchoua (Regional Delegate for Public Health of the West Region of Cameroon) for their support and advice.

## Supporting information caption

**Supplementary table 1.** Consolidated criteria for reporting qualitative studies (COREQ): 32-item checklist for interviews and focus groups. How just and fair are consent and debriefing for caesarean sections in Cameroon? An exploration of women’s and providers’ perspectives from 20 twenty hospitals in West Cameroon. March – August 2024.

